# Genomic surveillance reveals the emergence of SARS-CoV-2 Lineage A from Islamabad Pakistan

**DOI:** 10.1101/2021.12.24.21268367

**Authors:** Massab Umair, Aamer Ikram, Zaira Rehman, Syed Adnan Haider, Nazish Badar, Muhammad Ammar, Qasim Ali, Abdul Ahad, Rana Suleman, Muhammad Salman

## Abstract

The lineage A of SARS-CoV-2 has been around the world since the start of the pandemic. In Pakistan the last case of lineage A was reported in April, 2021 since then no case has been reported. In November, 2021 during routine genomic surveillance at National Institute of Health we have found 07 cases of lineage A from Islamabad, Pakistan. The study reports two novel deletions in the spike glycoprotein. One 09 amino acid deletion (68-76 a.a) is found in the S1 subunit while another 10 amino acid deletion (679-688 a.a) observed at the junction of S1/S2 referred as furin cleavage site. The removal of furin cleavage site may result in impaired virus replication thus decreasing its pathogenesis. The actual impact of these two deletions on the virus replication and disease dynamics needs to be studied in detail. Moreover, the enhanced genomic surveillance will be required to track the spread of this lineage in other parts of the country.

## Introduction

The etiological agent of the COVID-19 pandemic, Severe Acute Respiratory Syndrome 2 (SARS-CoV-2), is spreading globally. Despite the presence of VOCs Alpha (B.1.1.7), Beta (B.1.351), Gamma (P.1), Delta (B.1.617.2), and Omicron (B.1.1.529), several other SARS-CoV-2 lineages are circulating, potentially increasing the number of SARS-CoV-2 cases [1-3]. As of December 21, 2021, SARS-CoV-2 has infected 275,836,908 people worldwide, resulting in 5,377,893 fatalities. In Pakistan, however, 1,291,737 individuals were infected, resulting in 28,882 deaths [4]. Moreover, Pakistan is hit by four waves: the first from May to July 2020, the second from October 2020 to January 2021, and the third from March to May 2021[5]. The fourth wave, triggered by the delta variant, commenced in July 2021 [6].

Considering the upsurge in cases of SARS-CoV-2, Pakistan’s government is especially concerned about the impact and control measures of the circulating variants, which have evolved rapidly over the last two years [7, 8]. Consequently, between November 26 and December 8, 2021, the National Institute of Health (NIH) Islamabad collected samples for testing COVID-19, for which PCR tests performed resulted in false negative results for the spike gene (69/70del) target. Phylogenetic analysis of the sample’s sequences obtained by whole genome sequencing indicated lineage A (S Clade). As of December 21, 2021, 11 cases of individuals affected by the A lineage in Pakistan are listed in GISAID, with the first sample detected on June 02, 2020. At least 85 nations and 36 US states have reported a total of 2,783 cases of the A lineage [9]. The US accounts for 27%, the UAE has 13%, China has 9%, Germany has 8%, and Japan has 5% of the lineage A cases [10].

Lineage A of SARS-CoV-2, which is still in circulation, is the root of the pandemic, subsequently divided into sublineages [11, 12]. Of note, Alpha and Omicron share two important deletions (“H69/V70”) of the spike gene with lineage A. These deletions are used as a marker in PCR tests [13]. Additionally, H69/V70del compensates for immune escape mutations that impair infectivity [14-16]. Hence, understanding the virus’s ongoing evolution, epidemiology, circulating lineages, as well as evaluating the effects of spike protein mutations on COVID-19 transmission and vaccine performance are all essential for COVID-19 mitigation and control, which can be assisted by genomic surveillance studies[17]. As a result, the current finding of lineage A (clade S) cases in Pakistan underscores the relevance of genomic surveillance studies in directing healthcare officials in COVID-19 management decisions.

## Materials and Methods

### Sample Collection and Sample Processing

The department of Virology at National Institute of Health is performing routine genomic surveillance of SARS-CoV-2. As part of routine surveillance process during November 24, to December 08, 2021 the National Institute of Health received the nasopharangyel samples from 800 COVID-19 suspected patients for SARS-CoV-2 testing. Following RNA extraction using KingFisher™ Flex Purification System (ThermoFisher Scientific, US), the samples were subjected to SARS-CoV-2 testing using TaqPath™ COVID-19 CE-IVD RT-PCR kit (ThermoFisher Scientific, Waltham, US). Out of 317 positive samples, 07 samples were found to be spike gene target failure (SGTF). These samples were further subjected to whole genome sequencing.

### Next Generation Sequencing

The Illumina DNA Prep Kit (Illumina, Inc, USA) was used to prepare the paired-end (2×150 bp) sequencing library according standard protocol. The prepared libraries were pooled and subjected to sequencing on Illumina platform, iSeq, using sequencing reagent, iSeq 100 i1 Reagent v2 (300-cycle) (Illumina, Inc, USA).

### Data Analysis

The quality of sequencing reads were assessed through FastQC tool (v0.11.9) [18]. The low quality low-quality base calls (< 30) and adapter sequences were removed using Trimmomatic (v0.39). The alignment of filtered reads were performed through Burrows-Wheeler Aligner’s (BWA, v0.7.17) using Wuhan-WHU-01 (GISAID ID: EPI_ISL_402125). The variant identification and consensus sequence generation was performed according to Centers for Disease Control and Prevention (CDC, USA) guidelines. All consensus sequences were assigned to lineages by Pangolin v.3.1.16 (PangoLEARN v3/25.11.2021).

### Phylogenetic Analysis

For the phylogenetic analyses of the Pakistani lineage A strains in comparison with the globally reported sequences of lineage A, pyhlogenetic analysis was performed using Ultrafast Sample placement on Existing tRee (UShER). The placement of study samples were performed using the updated version on December 20, 2021.

### Multiple sequence alignment and structure prediction

The multiple sequence alignment of all the study isolates were performed with the Wuhan-Hu-01 (GISAID ID: EPI_ISL_402125) using Clustal W. The homology model of spike glycoprotein was built through Modeller V9.2 using pdb ID:6VSB as template and 100 models were generated. These models were subjected to model evaluation using Ramachandran Plot and Q-mean score.

## Results

During November 24, to December 8, 2021, a total of 07 SARS-CoV-2 positive samples were found to be the SGTF which subsequently underwent whole genome sequencing. There were 05 males and 02 females with age range of 21-50 years. All the seven patients were from Islamabad region. The patients had no history of travel. The patients do not have any severe symptoms of disease.

All the 07 sequences belonged to lineage A. The detailed sequence analysis revealed the presence of two unusual large deletions in spike glycoprotein in 05 of the study isolates. A 27 nucleotide (21764-21791) deletion was observed in the genome encoding the 68-76 amino acids region (referred as site I in the manuscript) of spike glycoprotein. Another 30 nucleotide deletion (23597-23627) was observed in the furin cleavage site encoding the amino acids 679-688 (referred as site II in the manuscript). Deletion in these residues can abolish the furin cleavage site. The other two samples of lineage A harbors ambiguous nucleotides at these two site (**Figure 1**). Phylogenetic analysis revealed the sequences of lineage A belonged to the sequences from China reported in 2020 (**Figure 2**).

**Figure 1:**
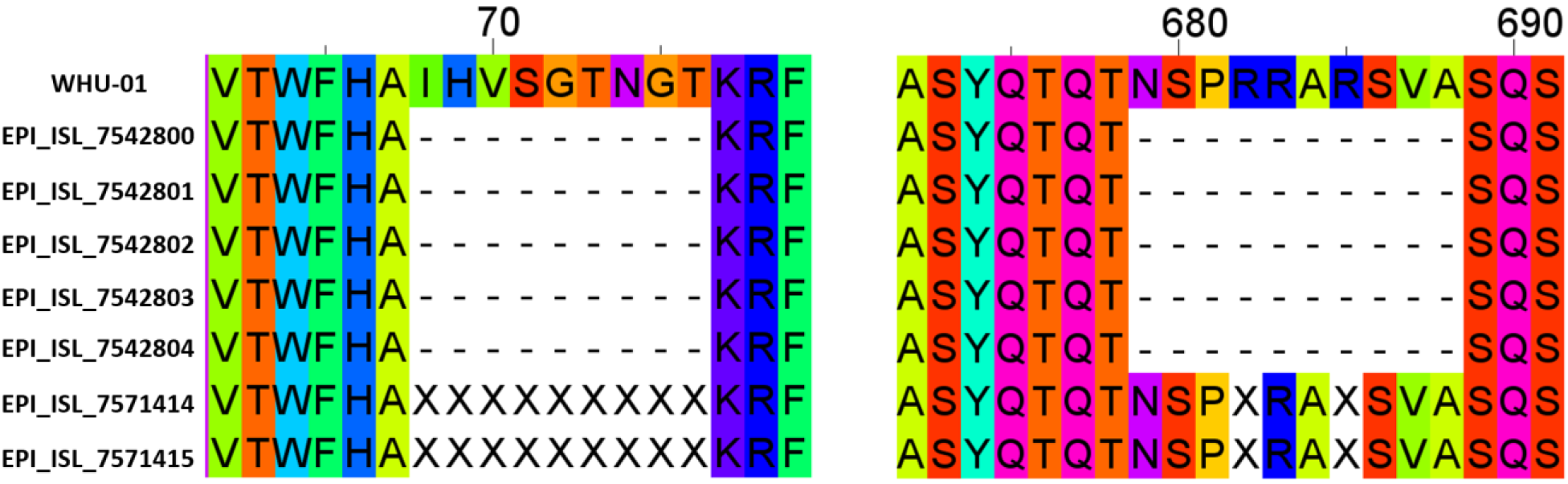
Portion of spike glycoprotein sequence alignment of seven isolates with Wuhan-01 as reference sequence. A 9 amino acid deletion was observed spanning the 68-76 amino acid long region (left panel) while a 10 amino acid deletion was observed spanning the 679-688 amino acid region (right panel).

**Figure 2:**
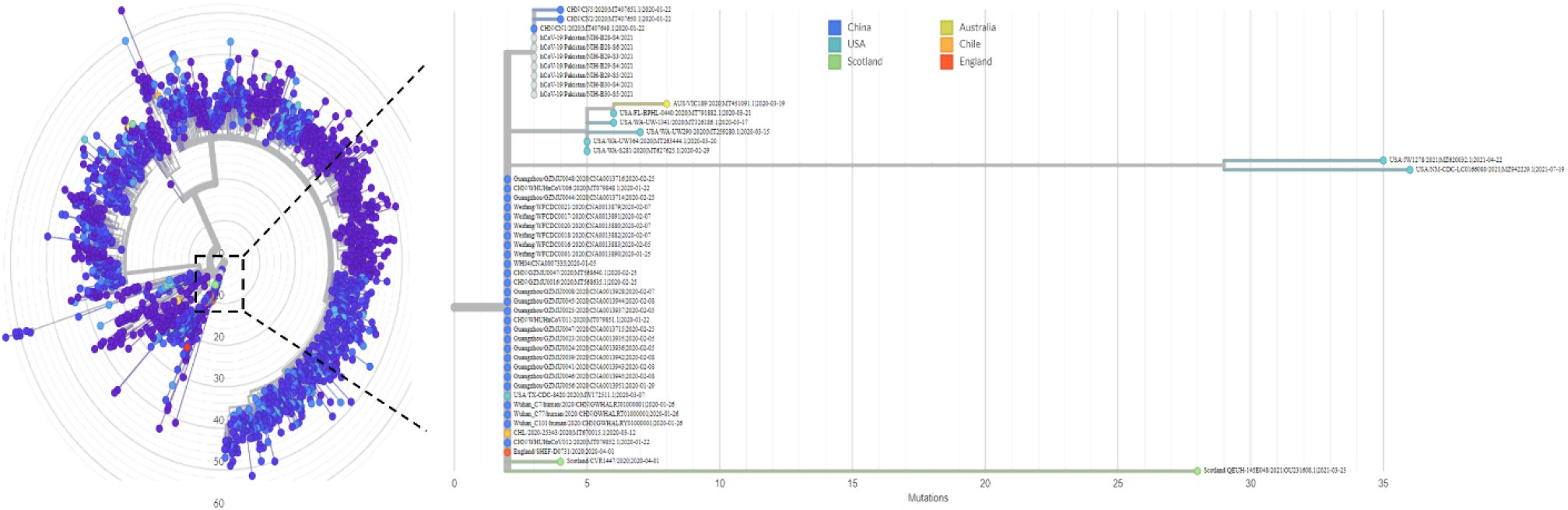
Phylogenetic analysis of study isolates. The Pakistani isolates are grey in color.

### Impact of 68-76 and 682-689del on structure of Spike glycoprotein

The structure of spike protein of wild type SARS-CoV-2 was superimposed on the spike structure with the deletions. The detailed structural analysis revealed that two (68-76 and 679-688) loops are missing in the lineage A spike protein (**Figure 3**). The structural comparison have also shown RMSD value of 1.7 Å from the native structure that further verified conformational changes in the structure.

**Figure 3:**
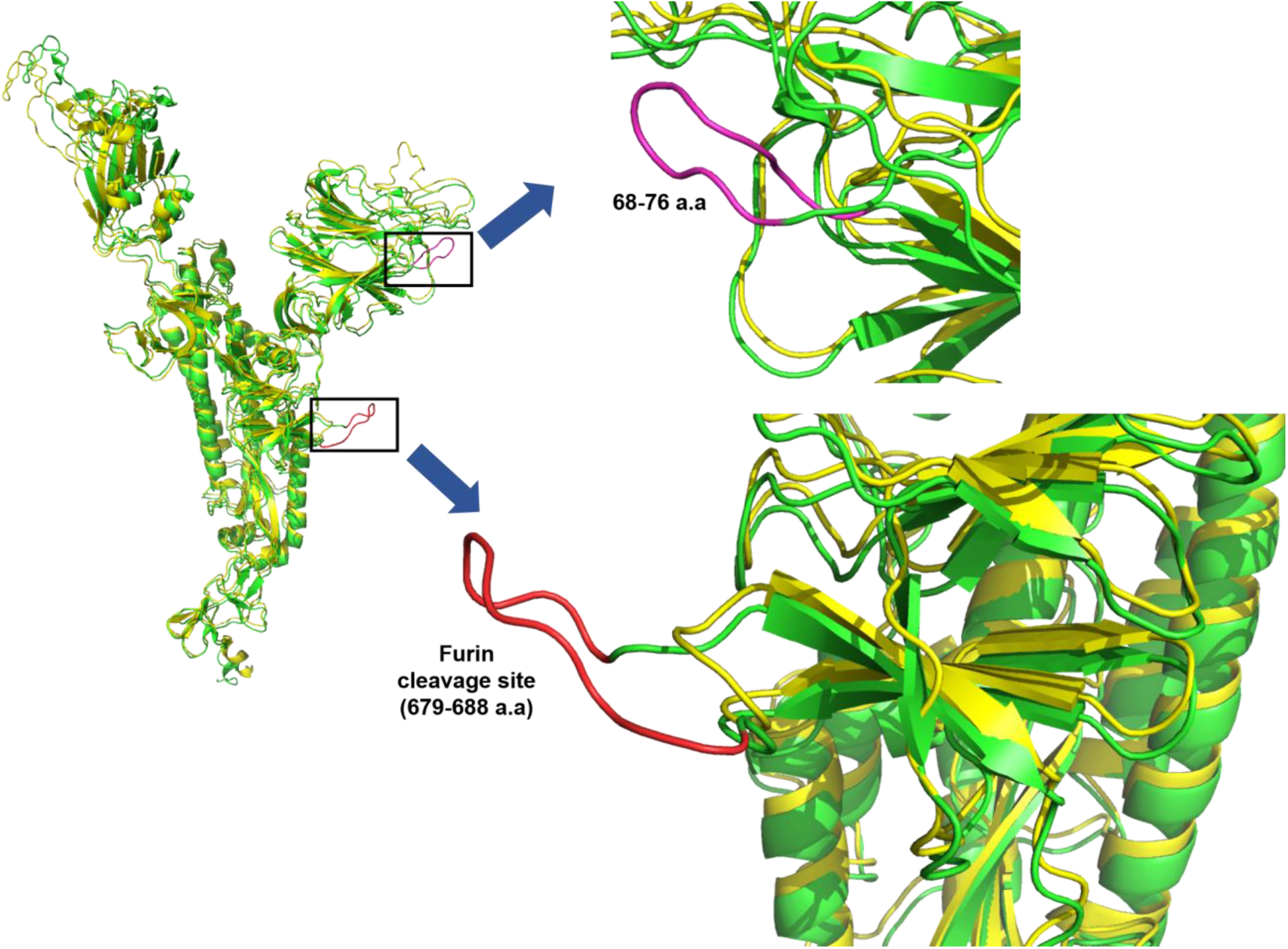
Superimposed structure of spike glycoprotein. The wild type spike glycoprotein is represented in green color while the structure of spike protein with deletions at site I and II is represented in yellow color. The 68-76 amino acid deletion in the N-terminal domain is shown in purple color (upper right panel) while 679-688 amino acids deletion is shown in red color (lower right panel).

## Discussion

This is the first study to report the emergence of lineage A with two large deletions (68-76 and 679-688del) in the spike protein from Pakistan. These deletions have to be seen in light of the functional significance. Typically the spike glycoprotein of SARS-Cov-2 has been proven to be involved in entry of virus into host cells by binding with the angiotensin-converting enzyme 2 (ACE2) [19, 20]. The spike protein is composed of two subunits, an N-terminal S1 subunit responsible for receptor binding and a C-terminal S2 subunit responsible for virus fusion with cell membrane [21, 22]. Previously, from Pakistan, the first three cases of lineage A have been reported in June, 2020. Later on in April, 2021 another case of lineage A was reported from Pakistan. Interestingly, the previously reported lineage A have no significant changes in the genome but the recently detected cases harbors two large deletions in the spike glycoprotein. According to globally reported data, till date there has been 2,776 isolates of Lineage A reported on GISAID (as of December 20, 2021). Out of these isolates only 21 sequences have been found to be having the 69-70del in the spike glycoprotein. This 69-70del is one of the characteristic deletion found in alpha variant and has also been observed in some of delta cases (n= 4896). Recently, the newly emerged omicron variant also harbors this deletion in the spike glycoprotein. The presence of 69-70del in different lineages of SARS-CoV-2 at different time points towards a typical temporal viral evolutionary trend. The first sequence of lineage A with the 69-70del was reported on May 4, 2020 in Madagascar, Africa while the most recent case was reported on June 28, 2021 in Italy. Interestingly, only two of the isolates (GISAID ID: EPI_ISL_2886419; EPI_ISL_2832924) from Germany in April, 2021 harbors the site I deletion (68-76 amino acids) in their genomes while these isolates devoid of the site II deletion. Instead of site II deletion these two isolates possess 676-680 amino acid deletion in the spike glycoprotein. This study has also identified 68-76 amino acid deletion in the S1 subunit. Previously the two amino acid deletion (69-70del) has been studied more extensively and found to have an impact on increasing infectivity as well as increasing the susceptibility to neutralising antibodies by a conformational change thus favoring a more open spike conformation [14]

The other major deletion identified in the studied samples spanned in the 679-688 amino acid region. Functionally, at the junction of S1 and S2, the specific sequence motif 682-689 amino acid region is recognized and cleaved by furin proteases during viral packaging thus promoting virus infectivity and pathogenicity. This site has been identified in MERS-CoV and SARS-CoV-2 however, it is absent in other coronaviruses of the same family [23, 24]. It has been reported previously, that deletion of furin cleavage site results in attenuated viral replication thus reducing the pathogenesis and ablating the disease. Thus presence of furin cleavage site is crucial for viral replication. The loss of furin cleavage site may also result in altered antibody neutralization profile [25-28].

The current study studied samples collected from the Capital Territory Region Islamabad where the positivity rate drop has dropped to <0.5% where now potential emergence of lineage A of SARS-CoV-2 with novel deletions in the spike glycoprotein is being noted. As the sequencing has been limited to a single city, in order to study the spread of this variant into different parts of the country, enhanced genomic surveillance will be required. Moreover, currently the country has increased vaccination rate with 27% population being fully vaccinated and 12% being partially vaccinated. Whether the emerging new lineages of SARS CoV2 can affect the already vaccinated individuals other than the non-vaccinated ones, requires a watchful eye to follow and therefore require a more robust genomic surveillance encompassing larger cohort and cities.

## Data Availability

All the sequences generated in the current study are submitted to the GISAID that are available at https://www.gisaid.org/login/ under the accession numbers: EPI_ISL_7542800-EPI_ISL_7542804, EPI_ISL_7571414-EPI_ISL_7571415.

